# Predictions, role of interventions and effects of a historic national lockdown in India’s response to the COVID-19 pandemic: data science call to arms

**DOI:** 10.1101/2020.04.15.20067256

**Authors:** Debashree Ray, Maxwell Salvatore, Rupam Bhattacharyya, Lili Wang, Shariq Mohammed, Soumik Purkayastha, Aritra Halder, Alexander Rix, Daniel Barker, Michael Kleinsasser, Yiwang Zhou, Peter Song, Debraj Bose, Mousumi Banerjee, Veerabhadran Baladandayuthapani, Parikshit Ghosh, Bhramar Mukherjee

**Affiliations:** Department of Epidemiology, Johns Hopkins University; Department of Biostatistics, Johns Hopkins University; Department of Biostatistics, University of Michigan; Center for Precision Health Data Science, University of Michigan; Department of Computational Medicine and Bioinformatics, University of Michigan; Department of Statistics, University of Connecticut; Department of Economics, Delhi School of Economics; Institute for Healthcare Policy and Innovation, University of Michigan

## Abstract

**Importance:** India has taken strong and early public health measures for arresting the spread of the COVID-19 epidemic. With only 536 COVID-19 cases and 11 fatalities, India – a democracy of 1.34 billion people – took the historic decision of a 21-day national lockdown on March 25. The lockdown was further extended to May 3rd, soon after the analysis of this paper was completed.

**Objective:** To study the short- and long-term impact of an initial 21-day lockdown on the total number of COVID-19 cases in India compared to other less severe non-pharmaceutical interventions using epidemiological forecasting models and Bayesian estimation algorithms; to compare effects of hypothetical durations of lockdown from an epidemiological perspective; to study alternative explanations for slower growth rate of the virus outbreak in India, including exploring the association of the number of cases and average monthly temperature; and finally, to outline the pivotal role of reliable and transparent data, reproducible data science methods, tools and products as we reopen the country and prepare for a post lock-down phase of the pandemic.

**Design, Setting, and Participants:** We use the daily data on the number of COVID-19 cases, of recovered and of deaths from March 1 until April 7, 2020 from the 2019 Novel Coronavirus Visual Dashboard operated by the Johns Hopkins University Center for Systems Science and Engineering (JHU CSSE). Additionally, we use COVID-19 incidence counts data from Kaggle and the monthly average temperature of major cities across the world from Wikipedia.

**Main Outcome and Measures:** The current time-series data on daily proportions of cases and removed (recovered and death combined) from India are analyzed using an extended version of the standard SIR (susceptible, infected, and removed) model. The eSIR model incorporates time-varying transmission rates that help us predict the effect of lockdown compared to other hypothetical interventions on the number of cases at future time points. A Markov Chain Monte Carlo implementation of this model provided predicted proportions of the cases at future time points along with credible intervals (CI).

**Results:** Our predicted cumulative number of COVID-19 cases in India on April 30 assuming a 1-week delay in people’s adherence to a 21-day lockdown (March 25 – April 14) and a gradual, moderate resumption of daily activities after April 14 is 9,181 with upper 95% CI of 72,245. In comparison, the predicted cumulative number of cases under “no intervention” and “social distancing and travel bans without lockdown” are 358 thousand and 46 thousand (upper 95% CI of nearly 2.3 million and 0.3 million) respectively. An effective lockdown can prevent roughly 343 thousand (upper 95% CI 1.8 million) and 2.4 million (upper 95% CI 38.4 million) COVID-19 cases nationwide compared to social distancing alone by May 15 and June 15, respectively. When comparing a 21-day lockdown with a hypothetical lockdown of longer duration, we find that 28-, 42-, and 56-day lockdowns can approximately prevent 238 thousand (upper 95% CI 2.3 million), 622 thousand (upper 95% CI 4.3 million), 781 thousand (upper 95% CI 4.6 million) cases by June 15, respectively. We find some suggestive evidence that the COVID-19 incidence rates worldwide are negatively associated with temperature in a crude unadjusted analysis with Pearson correlation estimates [95% confidence interval] between average monthly temperature and total monthly incidence around the world being −0.185 [−0.548, 0.236] for January, −0.110 [−0.362, 0.157] for February, and −**0.173 [−0.314, −0.026**] for March.

**Conclusions and Relevance:** The lockdown, if implemented correctly in the end, has a high chance of reducing the total number of COVID-19 cases in the short term, and buy India invaluable time to prepare its healthcare and disease monitoring system. Our analysis shows we need to have some measures of suppression in place after the lockdown for the best outcome. We cannot heavily rely on the hypothetical prevention governed by meteorological factors such as temperature based on current evidence. From an epidemiological perspective, a longer lockdown between 42-56 days is preferable. However, the lockdown comes at a tremendous price to social and economic health through a contagion process not dissimilar to that of the coronavirus itself. Data can play a defining role as we design post-lockdown testing, reopening and resource allocation strategies.

**Software:** Our contribution to data science includes an interactive and dynamic app (covind19.org) with short- and long-term projections updated daily that can help inform policy and practice related to COVID-19 in India. Anyone can visualize the observed data for India and create predictions under hypothetical scenarios with quantification of uncertainties. We make our prediction codes freely available (https://github.com/umich-cphds/cov-ind-19) for reproducible science and for other COVID-19 affected countries to use them for their prediction and data visualization work.

## Introduction

Four months since the first case of COVID-19 in Wuhan, China, the SARS-CoV-2 virus has engulfed the world and has been declared a global pandemic.^1^ The number of confirmed cases worldwide stands at a staggering 1,930,780 (as of 9:20 AM EST April 14, 2020, Microsoft bing coronavirus tracker^2^). Of these, 10,815 confirmed cases are from India (**Figure 1**), the world’s largest democracy with a population of 1.34 billion (compare China at 1.39 billion and USA at 325.7 million).^3^ India has been vigilant and wise in instituting the right public health interventions at the right time including sealing the borders with travel ban/canceling almost all visas, closing schools and colleges in certain states and diligently following up with community inspection of suspected/exposed cases with respect to adherence of quarantine recommendations (**Table 1**). On March 24, India took the historic decision of a 21-day national lockdown starting March 25, when it had reported only 536 COVID-19 cases and 11 fatalities. In the subsequent days we have seen a steady growth in the number of new cases and fatalities, with growth rates slower than other affected countries but in 21 days, the curve has not yet “turned the corner” or showed a steady decline in the number of newly diagnosed cases (**Figure 2**).

**Table 1.** Timeline of COVID-19 interventions in India.

| Date | Interventions |
| --- | --- |
| 3 March 2020 | ● India travel ban has been issued on four countries - China, South Korea, Italy, and Iran |
| 6 March 2020 | ● Union Health Ministry has issued advisory to avoid mass gatherings |
| 7 March 2020 | ● Agra Mayor has urged the Union government to close down historical monuments including Taj Mahal<br>● Kuwait suspends flights to India |
| 9 March 2020 | ● Qatar puts India on travel ban list |
| 10 March 2020 | ● Manipur has closed its border with Myanmar |
| 11 March 2020 | ● India suspends all visas and e-visas granted to nationals of France, Germany, and Spain on or before 11 March |
| 12 March 2020 | ● **WHO declared the COVID-19 outbreak as 'pandemic'**<br>● India suspends all visas with the exception of those for diplomatic, UN, or international bodies, official and employment purposes until 15 April<br>● India reports 1st death |
| 13 March 2020 | ● India reports 2nd death<br>● Several academic institutions (e.g., JNU, IIT, IIM) cancel classes/convocations, some closed hostels |
| 16 March 2020 | ● Centre proposes social distancing measures until 31 March<br>● India bans passengers from EU countries, UK, and Turkey until the end of March<br>● Central government recommends the closure of educational institutions until 31 March |
| 17 March 2020 | ● Taj Mahal will be shut down until 31 March; ASI closes 3,000 monuments and 200 museums<br>● Mandatory quarantine of passengers coming from UAE, Qatar, Oman, and Kuwait<br>● **India is heading towards a countrywide lockdown mode** |
| 19 March 2020 | ● India halts all incoming commercial international flights for 1 week<br>● Some state governments ban public transportation<br>● Prime Minister urges people of India to observe self-imposed curfew on March 22 |
| 20 March 2020 | ● Maharashtra announce lockdown in Mumbai, Nagpur and Pune<br>● JNU orders students to vacate hostels |
| 21 March 2020 | ● Private labs can conduct COVID-19 tests, says Maharashtra government |
|  | ● Rajasthan Government declares lockdown until March 31 |
| 22 March 2020 | ● 12 states including Telangana and Delhi announce lockdown until March 31<br>● International commercial passenger flights are not allowed to land in India from today for one week<br>● Railways suspends all train services until March 31 |
| 23 March 2020 | ● **Centre has ordered all states in India to impose lockdown**<br>● Legal action will be initiated against people violating lockdown measures |
| 24 March 2020 | ● **Indian Prime Minister Narendra Modi has announced lockdown for 21 days as country records 552 coronavirus cases and ten deaths** |
| 28 March 2020 | ● Indian government unveiled stimulus package to help those hit by 21-day lockdown<br>● Focus right now is to construct COVID-19 hospitals, sample testing, contact-tracing and social distancing: Union Health ministry |
| 2 April 2020 | ● Common exit strategy necessary for “staggered” relaxations after lockdown period ends: PM Modi tells CMs |
| 6 April 2020 | ● PM tells union ministers to prepare a graded plan to gradually open departments that are not COVID-19 hotspots |
| 8 April 2020 | ● PM Modi, CMs to decide on lockdown extension on 11 April |
| 9 April 2020 | ● Odisha extends lockdown until 30 April and becomes first Indian state to do so |

Source: https://www.pharmaceutical-technology.com/news/india-covid-19-coronavirus-updates-status-by-state/

**Figure 1.**
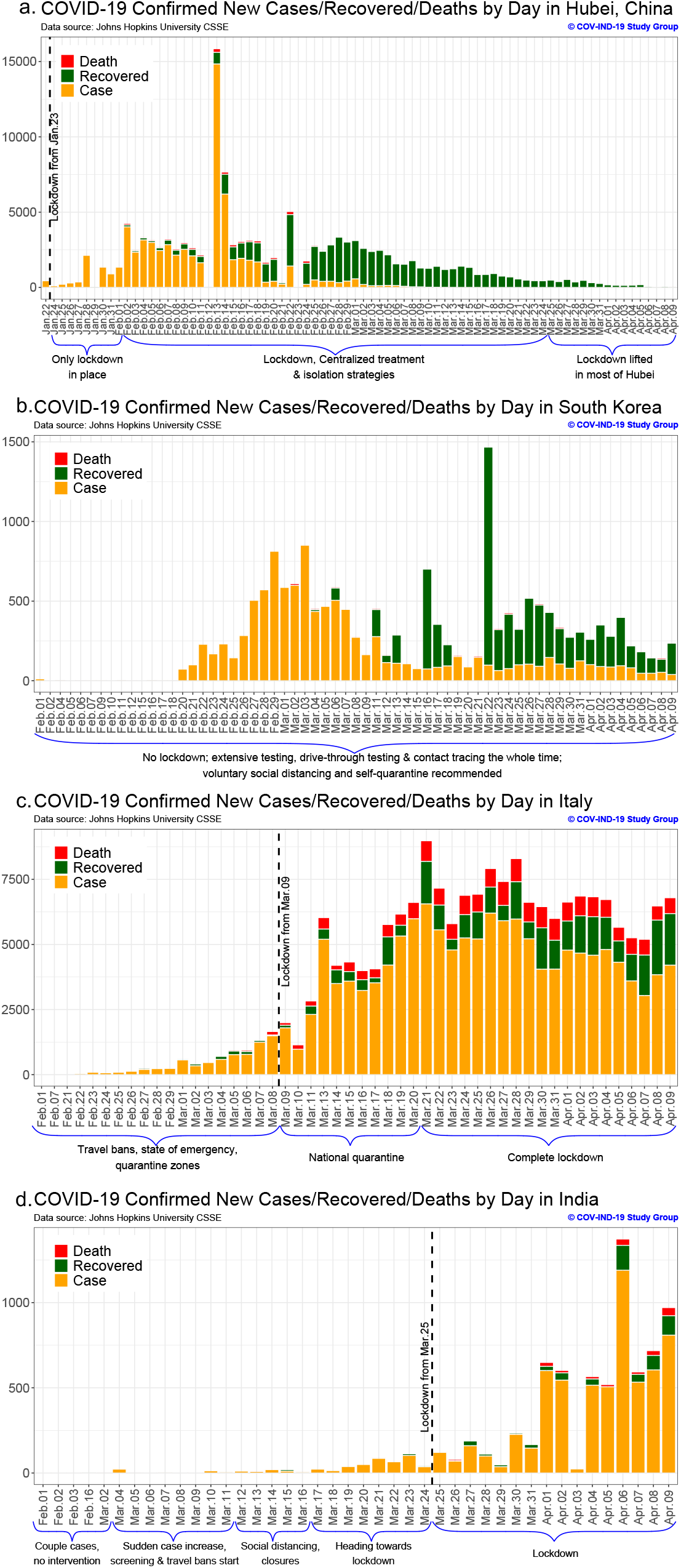
Description of the cases, recovered and fatalities in India with landmark policy/recommendations.

**Figure 2.**
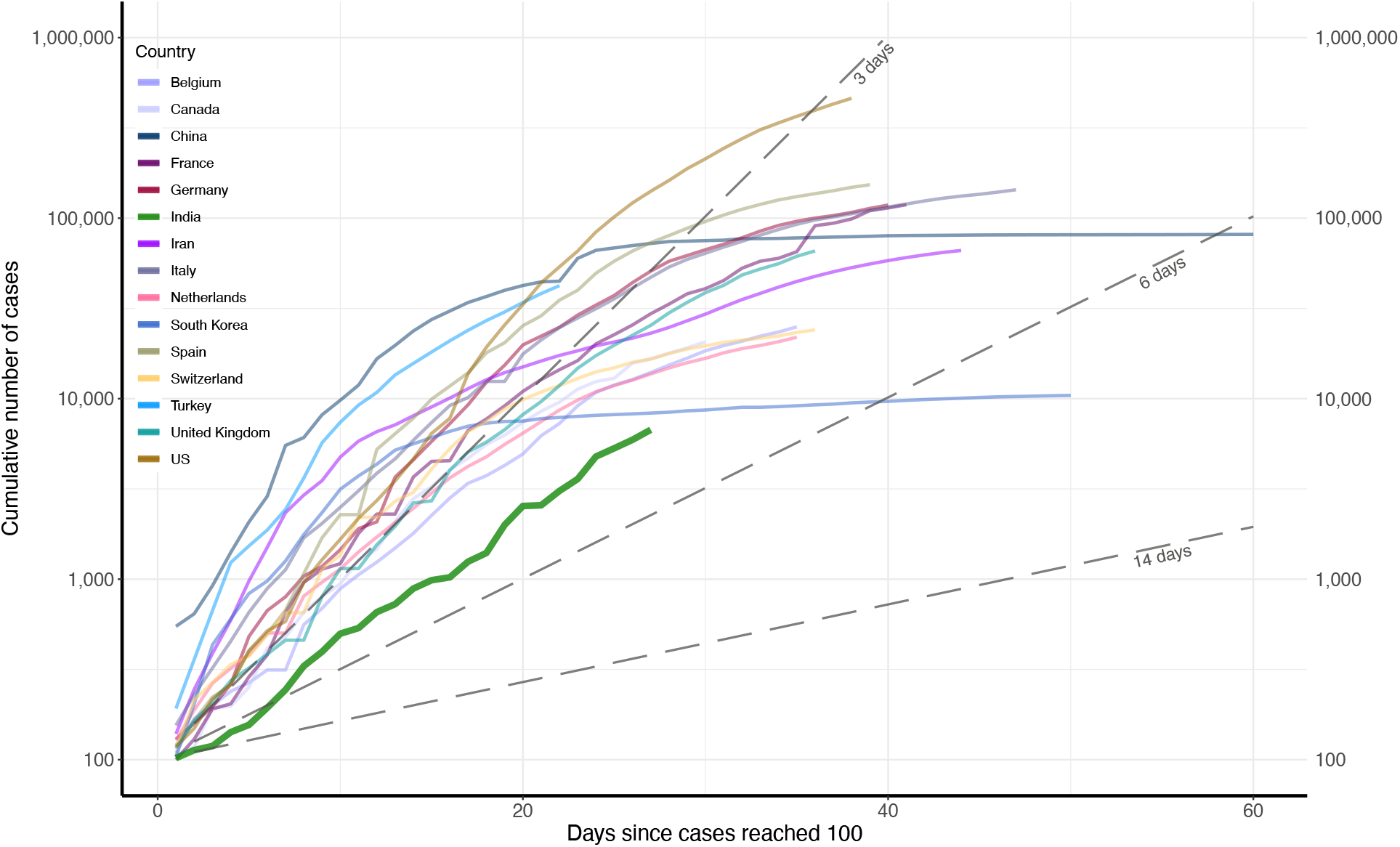
Early phase of the epidemic and daily growth in cumulative COVID-19 case counts in India compared to other countries affected by the pandemic using data through April 9.

While India seems to have done relatively well in controlling the number of confirmed cases compared to other countries in the early phase of the pandemic (**Figure 2**), there is a critical missing or unknown component in this assessment: “The number of truly affected cases,” which depends on the extent of testing, the accuracy of the test results and, in particular, the frequency and scale of testing of asymptomatic cases who may have been exposed. The frequency of testing has been low in India. According to the Indian Council of Medical Research (ICMR), only 130,792 subjects have been tested as of April 9.^4^ When there is no approved vaccine or drug for treating COVID-19, entering phase 2 or phase 3 of escalation will have devastating consequences on both the already overstretched healthcare system of India, and India’s large at-risk sub-populations (**Supplementary Table 1**). As seen for other countries like the US or Italy, COVID-19 enters gradually and then explodes suddenly. We provide a table listing other highly affected countries along with their first reported case, initial interventions, crude fatality rates, and active case counts in **Supplementary Table 2** for reference.

In this article, we take a data-driven approach to explore five extremely time-sensitive and important questions that India faces today in light of the COVID-19 outbreak and the national lockdown: (a) How many cases can India expect at the end of the lockdown period? (b) When will the curve in India reach its apex and will the number of cases go back up after lockdown is lifted? (c) Can summer temperatures thwart the outbreak in India? (d) How can the government and the people of India prepare for this crisis during and after the lockdown? (e) How critical is it to have reliable data, data science methods and tools as we envision a long-term strategy during and after the lockdown?

This work is the result of the collective public health conscience of a group of interdisciplinary researchers in different parts of the US and in India. We convened virtually after being quarantined in our homes with alternating waves of fear and inspiration surrounding us. We decided to channel our collective energy to study the defining public health and economic crisis of our time and use our data science expertise to search for answers and solutions that can help COVID-19 related policymaking in India. This is our contribution and public service as data scientists. Our data science product includes two articles on Medium <u>pre</u>^5^ and <u>post</u>^6^ lockdown announcement, providing critical information for policymakers (Reuters,^7^ Times of India,^8^ The Guardian,^9^ The Economic Times^10^) and an <u>interactive app</u> that daily updates forecasts as new case counts are coming in, and publicly available codes for reproducible research.

## Methods

### Study design and data sources

We used the current daily data on number of COVID-19 cases, recoveries and deaths in India to predict the number of cases at any given time.^11^ We obtained the data (up to April 7) from the 2019 Novel Coronavirus Visual Dashboard operated by the Johns Hopkins University Center for Systems Science and Engineering (JHU CSSE).^12,13^ For our temperature analysis, these counts were aggregated to a month-level for each country, that is, we look at the total number of new cases in the months of January, February and March for each country. We obtained the monthly average temperature for major cities in the countries with COVID-19 outbreak from Wikipedia.^14^

### Statistical model for predictions

We analyzed the data from India with standard epidemiologic tools of modeling disease transmission and estimating the theoretical number of cases at any time. One such epidemiologic model is the susceptible-infected-removed (SIR) model, which is guided by a set of differential equations relating the number of susceptible people, the number of infected people (cases) and the number of people who have been removed (either recovered or dead) at any given time. Recently, this standard SIR model was extended to incorporate time-varying transmission rates or time-varying quarantine protocols and is known as the eSIR model.^11^ When using the eSIR model with time-varying disease transmission rate, it can depict a series of time-varying changes caused by either external variation like government-initiated macro isolation measures, community-level protective measures and environment changes, or internal variations like mutations and evolutions of the pathogen. The R package for implementing this general model for understanding disease dynamics is publicly available at https://github.com/lilywang1988/eSIR.

To implement the eSIR model, a Bayesian hierarchical framework is assumed where the proportions of infected and the removed people are modeled using a Beta-Dirichlet state-space model while a latent Dirichlet distribution is assumed for the underlying unknown prevalence of the three states. Priors for the basic reproductive number R_0_, disease removal rate (consequently, the transmission rate) and the underlying unobserved prevalence of the susceptible, infected and removed states at the starting time are considered. Using the current time series data on the proportions of infected and the removed people, a Markov chain Monte Carlo implementation of this Bayesian model provides not only posterior estimation on parameters and prevalence of all the three compartments in the SIR model, but also predicted proportions of the infected and the removed people at future time point. The posterior mean estimates of the unobserved prevalence at both observed as well as future time points come along with 95% credible intervals (CI). To get predicted case-counts from the predicted prevalence, we used 1.34 billion as the population of India, thus treating the country as a homogeneous system for the outbreak.^15^

### Parameter choices for short-term forecasts

We made projections of the cumulative number of cases over a time horizon to assess the short-term impact of lockdown as well as the long-term impact of lockdown and post-lockdown activities. For the short-term forecast on April 30, we assumed lockdown is implemented until April 14 with either a 1-or a 2-week delay in people’s adherence/compliance to lockdown restrictions. We compared these projections with two hypothetical scenarios: (A) no non-pharmaceutical intervention (i.e., a constant disease transmission rate over time since the first case was reported in India), (B) a moderate intervention with social distancing and travel bans only (i.e., a decreased transmission rate compared to no intervention). For the no intervention and the moderate intervention scenarios, we chose the transmission rate and the removal rate such that the means for the prior distribution of the basic reproductive number R_0_ (the expected number of cases generated by one infected person assuming that the whole population is susceptible) are 2.0 and 1.5 respectively [the change in R_0_ was created based on what we saw in Wuhan^16^]. The value of 2.0 was estimated based on the early phase data in India. For the current scenario of lockdown, our chosen mean for R_0_ prior starts with 2.0 during the period of no intervention, drops to 1.5 during the period of moderate intervention, and further drops to 0.8 during the 21-day lockdown period, and moves back up to 1.5 after the lockdown ends as described in **Figure 3** (assuming a gradual, moderate resumption of daily activities).

**Figure 3.**
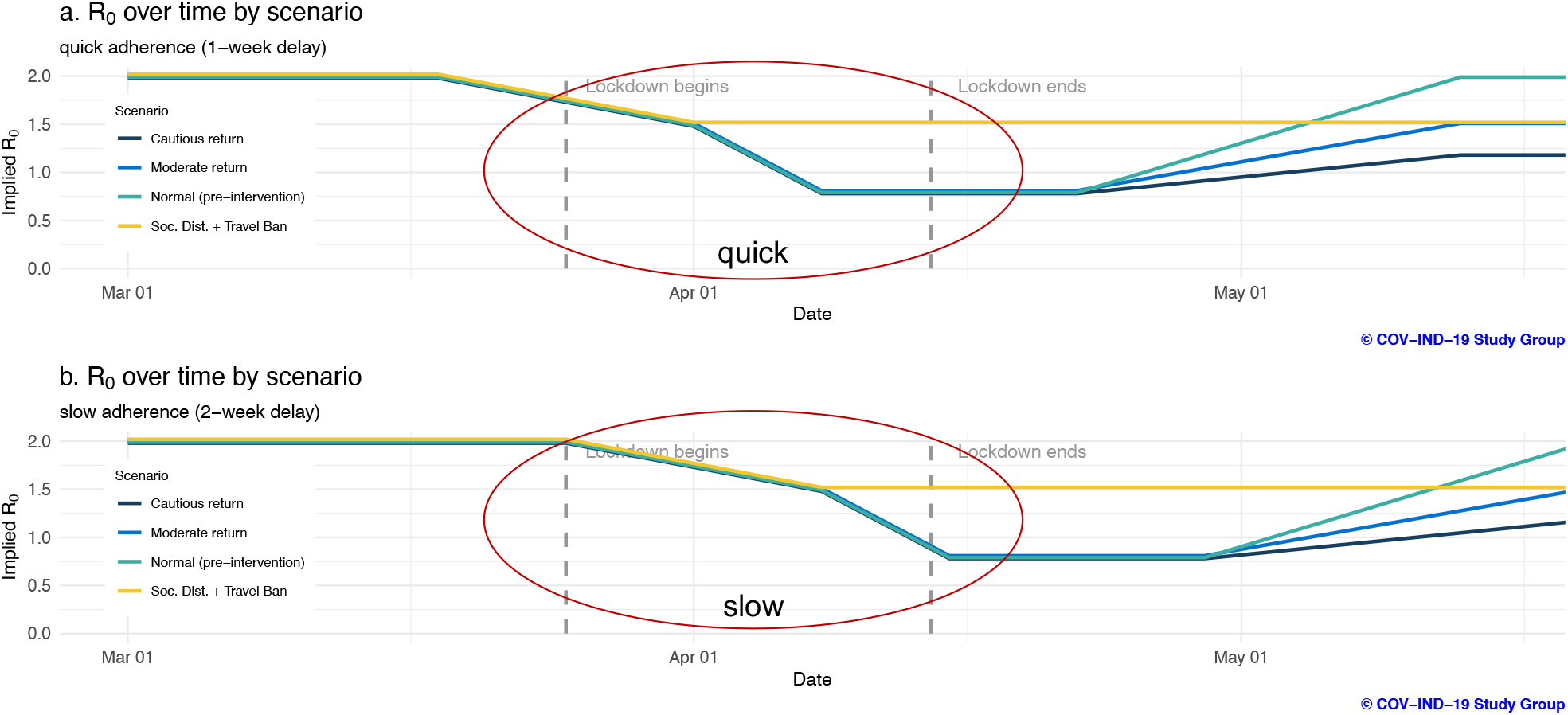
Implied R_0_ schedules corresponding to quick and slow adherence for the hypothetical scenarios.

### Parameter choices for long-term forecasts

For the longer-term forecast until June 15, we considered three hypothetical post-lockdown scenarios: (i) people return to normal activities due to the urgent desire for reconnecting after lockdown; (ii) people return to moderate activities as they did during the period with social distancing and travel ban intervention; and (iii) people make a cautious return out of fear for the coronavirus and partake in subdued activities. For these three scenarios, we assume mean for R_0_ prior moves back up from 0.8 to 2.0, 1.5 and 1.2 respectively three weeks after lockdown ends on April 14. We compared these post-lockdown scenarios with another hypothetical scenario involving perpetual social distancing and travel ban only without any lockdown (we fixed the mean for R_0_ prior at 1.5 over the entire intervention interval). The changes to R_0_ values across our simulation scenarios are depicted in **Figure 3**.

### Parameter choices for duration of lockdown analysis

To assess the long-term impact of lockdown duration, we considered four scenarios: 21-, 28-, 42-, and 56-day lockdown periods. In all scenarios, we assume mean for R_0_ prior remains at 0.8 for the duration of the lockdown and returns to 1.5 three weeks after the lockdown period ends (analogous to the “moderate return” scenario). The changes to R_0_ values across our simulation scenarios are depicted in **Supplementary Figure 4**.

### Statistical model for temperature analysis

There are many hypotheses regarding the slow growth rate of COVID-19 cases in many countries, particularly low- and middle-income countries (LMICs). Some of these hypotheses include the use of Bacille Calmette Guerin (BCG) vaccine,^17^ younger population,^18^ high daily temperature,^19^ use of anti-malarials^20^ and host genetics.^21^ Here, we only explore the temperature hypothesis related to COVID-19 incidence. We assessed any correlation between country-wise average monthly temperature and total incidence of COVID-19. The monthly average temperature for major cities across the world was used to compute the monthly average temperature for each country experiencing COVID-19 outbreak by averaging across the major cities within a country. Missing data for average temperature for certain countries was manually appended from www.weather-atlas.com.^22^ We computed the Pearson correlation coefficient, *r*, between the average monthly temperature and total monthly incidence during each month of January, February and March. We used the Fisher’s z-transformation to compute 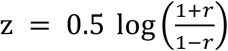. The standard deviation of *z*, which is known under certain normality assumptions, is used to construct a 95% confidence interval for *z*. The inverse transform of *z* is then used to obtain the 95% confidence interval for the correlation *r*. All calculations were carried out in the *RStudio* platform.

### Open-source software

We are committed to data transparency and reproducible research. Daily updates of our India projections, based on cases, recovered and deaths reported the day before by covid19india.org,^23^ a crowd-sourced database using state bulletins and official handles, can be found in our interactive and dynamic Shiny app (covind19.org). Apart from the scenarios described in this article, anyone can create predictions under other hypothetical scenarios with quantification of uncertainties. Open source codes behind this app are available at https://github.com/umich-cphds/cov-ind-19.

## Results

### Short-term forecast of cumulative case counts in India

Under national lockdown (March 25 – April 14), our predicted cumulative number of COVID-19 cases in India on April 30 are 9,181 and 11,626 (upper 95% CI of 72,245 and 84,245) assuming a 1-or 2-week delay (i.e., either a quick or a slow adherence), respectively, in people’s adherence to lockdown restrictions and a gradual, moderate resumption of daily activities post-lockdown (**Figure 4, Supplementary Figure 1**). In comparison, the predicted cumulative number of cases under “no intervention” and the “intervention involving social distancing and travel bans without lockdown” are 358 thousand and 46 thousand (upper 95% CI of nearly 2.3 million and 0.3 million) respectively. We are reporting only the upper credible limit here and elsewhere since the lower credible limits are very close to 0 due to the large uncertainty in our predictions arising from many unknowns. We also believe that our **point estimates are at best underestimates** due to potential under-reporting of case-counts and our model not taking into account the population density, age-sex and contact network structure of the whole nation. Increase in testing and community transmission may lead to a spike in a single day and that may shift the projection curve significantly upward. Regardless of the exact numbers it is clear that, the 21-day lockdown will likely have a strong effect on reducing the predicted number of cases in the short term.

**Figure 4.**
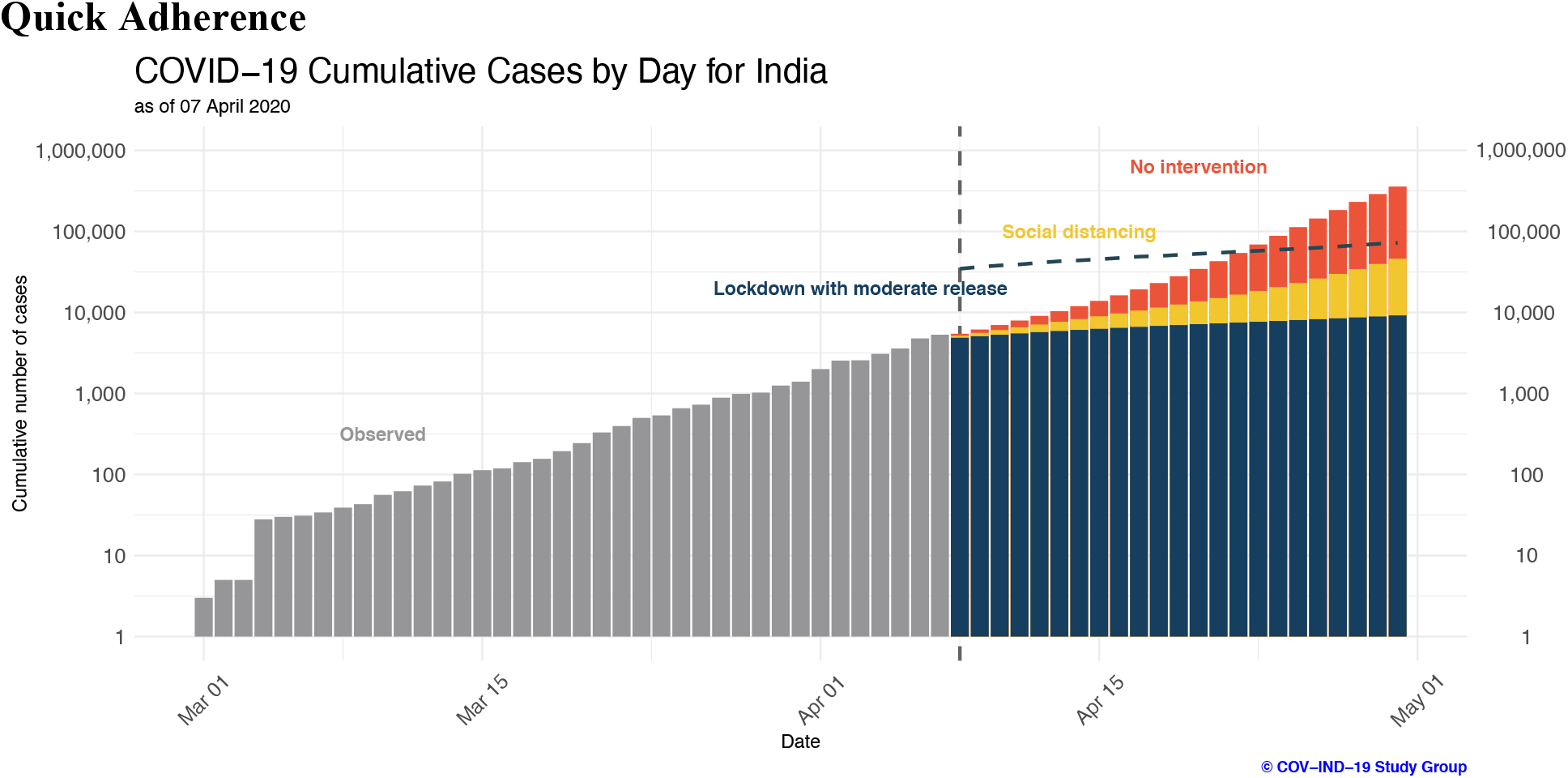
Short-term daily growth in cumulative case counts in India assuming a 1-week delay in people’s adherence to restrictions. Observed data are shown for days up to April 7. Predicted future case counts for April 8 until April 30 are based on observed data until April 7 using the eSIR model. Corresponding graph following a 2-week delay schedule can be found in **Supplementary Figure 1**.

### Long-term impact of lockdown on the outbreak in India

We took a close look at what might be coming in the next few months, based on what we have seen in other countries and an epidemiological model that has been gainfully employed to assess the effect of interventions in Hubei province.^11^ We estimated that 26 (upper 95% CI 135) and 177 (upper 95% CI 2869) cases *per 100,000* are avoided by May 15 and June 15, respectively, by instituting a 21-day lockdown with a 1-week delay in people’s adherence and a cautious release compared to perpetual social distancing and travel ban (without lockdown) (**Figure 5**). This boils down to **preventing roughly 343 thousand** (upper 95% CI 1.8 million) and **2.4 million** (upper 95% CI 38.4 million) COVID-19 cases *nationwide* by May 15 and June 15, respectively.

**Figure 5.**
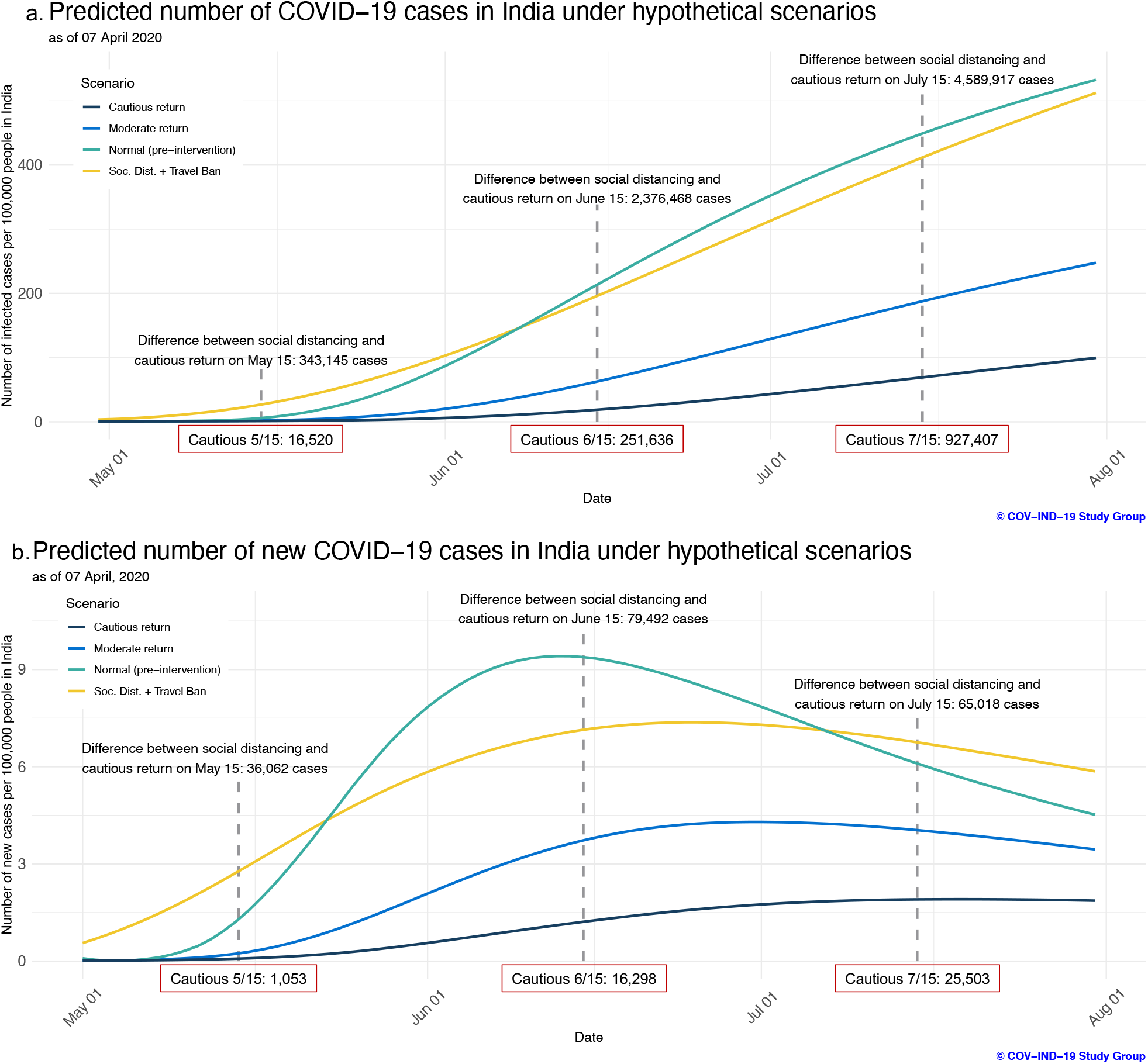
Long-term daily growth in case counts in India per 100,000 people assuming a 1-week delay and how that is affected by different non-pharmaceutical intervention strategies. Predicted cumulative (a) and incident (b) case counts from April 30 to July 31 from the eSIR model are shown, based on observed data until April 7. Corresponding plots for slow adherence are in **Supplementary Figure 2**.

Without some measures of suppression after lockdown is lifted, the impact of lockdown in bringing down the case-counts (the now ubiquitous term, “flattening the curve”) can be negated by as early as the first week of June. In fact, in **Figure 5a**, the pre-intervention (“normal”) curve first passes the social distancing and travel ban curve on June 8. In particular, if people immediately go back to pre-intervention (“normal”) activities post-lockdown, a surge in the predicted case-counts is expected in the long-term beyond what we would have seen if there were only social distancing and travel ban measures without lockdown (7.1 million when post-lockdown activity returns to pre-intervention levels vs. 6.9 million under social distancing and travel ban without a lockdown period on July 31; **Figure 5**). We estimated that 195 (upper 95% CI 3494) and 380 (upper 95% CI 6463) cases *per 100,000* are avoided by June 15 and July 15 respectively if people are cautious in their activities post-lockdown compared to the scenario where people return to normal pre-intervention activities. Long-term forecasting under slow adherence (2-week delay) can be seen in **Supplementary Figure 2**.

### Optimal duration of lockdown in India

We took the quick adherence epidemiological models and compared the 21-day lockdown with hypothetical 28-, 42-, and 56-day lockdown scenarios (**Figure 6**). When comparing a 21-day lockdown with a hypothetical lockdown of longer duration, we find that 28-, 42-, and 56-day lockdowns can approximately prevent 238 thousand (upper 95% CI 2.3 million), 622 thousand (upper 95% CI 4.3 million), 781 thousand (upper 95% CI 4.6 million) cases by June 15, respectively. A 28-day lockdown does not appear to have a significant impact on cumulative case counts when compared to a 21-day lockdown. However, purely from an epidemiologic perspective, there appears to be some evidence that suggests a 42-or 56-day lockdown would have a more meaningful impact on reducing cumulative COVID-19 case counts in India. We note that longer lockdown periods are accompanied by increasing costs to individuals - notably economic - and must be considered. Our models suggest that some form of post-lockdown suppression (e.g., extension of social distancing measures, limits of gathering size, etc.) is necessary to observe long-term benefits of the lockdown period.

**Figure 6.**
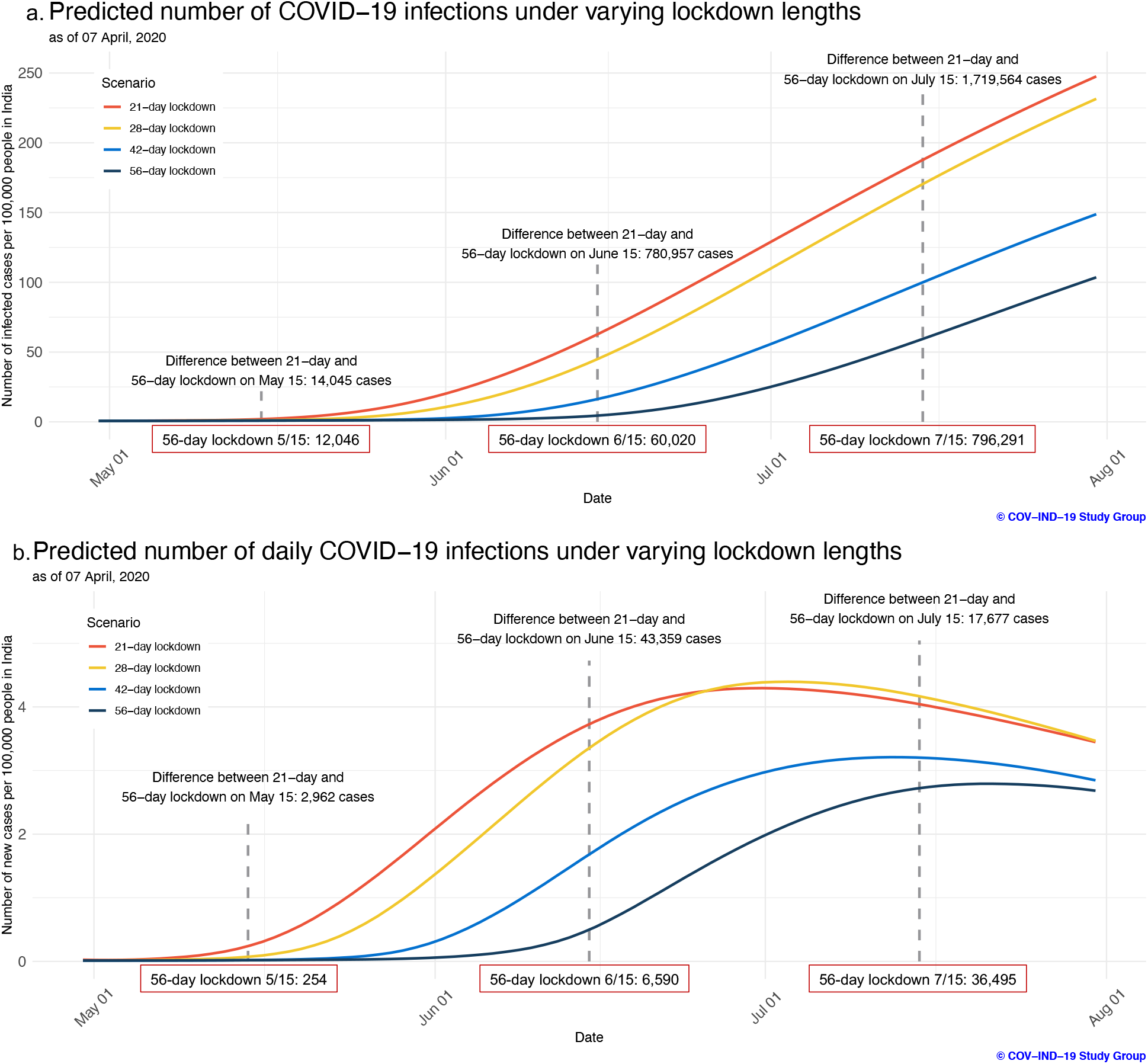
Cumulative (a) and incidence (b) graphs for forecasting models assuming a 1-week delay under 21-, 28-, 42-, and 56-day lockdown scenarios using observed data through April 7. Corresponding plots for slow adherence are in **Supplementary Figure 3**.

**Figure 7.**
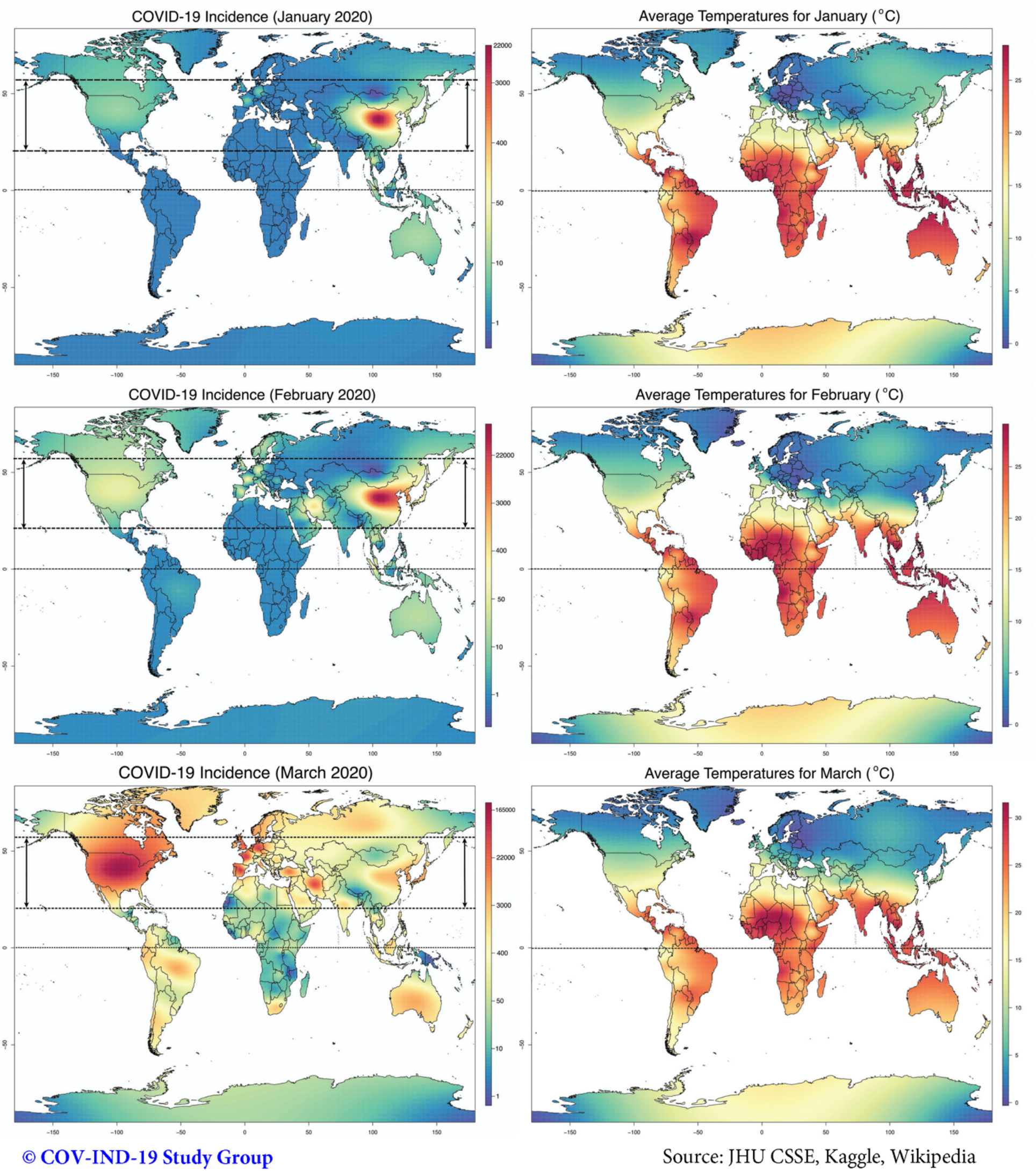
*Left*: Country-wise total monthly incidence of COVID-19 in the months of January, February and March. The horizontal lines approximately indicate the equator, the tropic of cancer and the 60N latitude. *Right*: Average monthly temperature (in C) during the months of January, February and March. These maps were created by smoothing the counts as well as the average monthly temperature across geographical locations by spatial interpolation.

Lockdown duration study under the slow adherence (2-week delay) scenario can be found in **Supplementary Figure 3**.

### Sensitivity analyses

We did explore some alternative assumptions and conducted thorough sensitivity analysis before settling on the models presented above. In one example, we assumed that there are actually 10 times the number of reported cases to date to reflect potential underreporting of cases due to lack of testing. In another scenario, we assumed these cases occurred in metropolitan areas to reflect a potential intensification of case clustering. In yet a third scenario, we hypothesized that R_0_ prior starts with 2.5 instead of 2.0 (i.e., a single infected individual would infect 2.5 susceptible individuals, on average, instead of 2). These scenarios did not appreciably change our conclusions in broad qualitative terms, though the exact quantitative projections are quite sensitive to such choices. Across these scenarios, the projected total number of infected cases by the entire first phase of the pandemic **varied between 2–15%** of the population, again showing the significant variability in these numbers. The estimates we present here may appear conservative and are at best underestimates, and, in all cases, our confidence in these projections decreases markedly the farther into the future we try to forecast. It is extremely important to update these models as new data arise.

### COVID-19 case counts and daily temperature

Spatial plots for the average monthly temperatures accompanied by total monthly incidence across all countries from January through March indicate a suggestive pattern of increase in community spread across cities and regions specifically along narrow north east-west directions (**Figure 6**). Countries in these regions consistently exhibit similar weather patterns. However, in the context of India, a gradual rise in the number of cases is observed starting from January through March.

The estimates and the 95% confidence interval for the correlation coefficient for January was - 0.185 [−0.548, 0.236] with 24 countries having non-zero incidence, for February was −0.110 [−0.362, 0.157] with 56 countries having non-zero incidence, and for March was −**0.173 [−0.314, −0.026]** with 175 countries having non-zero incidence. Although the estimates were negative, the 95% confidence intervals either include zero or the upper limit is close to 0, indicating weak evidence for any claim of negative association between case counts and daily temperature. Any such affirmation will require further data and investigation that accounts for many possible sources of confounding.

## Discussion

Our projections using current daily data on case counts until April 7 in India show that the lockdown, if implemented correctly in the end, has a high chance of reducing the number of COVID-19 cases in the short term and buy India invaluable time to prepare its healthcare and disease monitoring system. In the long-term, we need to have some measures of suppression in place after the lockdown is lifted to prevent a massive surge in the number of cases that can quickly overwhelm an already over-stretched Indian healthcare system resulting in increased fatalities. Specific vulnerable populations will be at higher risk of severity and fatality from COVID-19 infection: older persons and persons with pre-existing medical conditions (e.g., high blood pressure, heart disease, lung disease, cancer, diabetes, immunocompromised persons).^24,25^ **Supplementary Table 1** provides a description of the approximate number of individuals in these high-risk categories in India. Beyond the fragile population characterized by health and economic indicators, we have to remember that healthcare workers and first responders at the front line of this pandemic are amongst the most vulnerable.^16^

It is important to note that a massive surge in the number of cases can quickly overwhelm an already over-stretched Indian healthcare system. The estimated capacity of hospital beds in India is 70 per 100,000,^26^ which is an upper bound on treatment capacity. Given an average occupancy rate of 75%, only a quarter of these are available.^27^ Moreover, critically ill COVID-19 patients (about 5-10% of those infected) will require ICU beds and ventilator support. India has only 35– 58 thousand ICU beds with very high occupancy rates and at most 1 ventilator per 2 ICU beds.^28^ From a purely public health perspective, this analysis shows the impact and necessity of lockdown and subsequent measures of suppression after lockdown is lifted. All the people in India, regardless of their vulnerability to COVID-19, need to adhere to the public health guidelines issued by the Ministry of Health and Family Welfare in India, and continue to be cautious in their post-lockdown activities to guarantee a long-term benefit of the national lockdown.

Currently, there are many hypotheses regarding differential COVID-19 infection rates and mortality rates across countries. One such hypothesis is that the BCG vaccine – developed a century back for tuberculosis – has a protective effect on the prevalence of COVID-19 and related mortality.^29^ A recent pre-print found COVID-19 attributable mortality in countries with BCG policy is 6 times lower than those without a BCG policy in an ecological analysis, after accounting for country-specific confounders like economic status, percentage elderly (those aged ≥65 years) in the population, and relative position of each country along the epidemic trajectory.^17^ However, the authors caution against over-interpretation of this negative association between BCG use and COVID-19 due to limitations of country-level analysis and many sources of unmeasured confounders. Another hypothesis is that much like the flu virus, summer temperatures will thwart the COVID-19 outbreak. Our analysis, based on current data, suggests we cannot rely on the hypothetical prevention (with inconclusive evidence) governed by meteorological factors and need public health actions, regardless of the seasonal weather.

The management of this COVID-19 crisis requires strong partnership of the government, the scientific community, the health care providers and all citizens of India (and all global citizens). Long term surveillance and management of COVID-19 crisis is needed with not just public health in mind but also to take care of the economic, social, and psychological trauma that it will leave on the people. Reviving the economy will be critical in the coming months. Below we recommend some healthcare, social and economic reforms that can counter the negative impact of severe public health interventions, some of which India has already begun to make progress.

### Healthcare recommendations

a. Aggressively increase the number of tests administered daily as there are often asymptomatic cases who are spreading the infection without knowing. It is of utmost importance that India adopt widespread testing to identify and isolate the infected. RT-PCR diagnostic test can provide reliable and faster diagnosis of the SARS-CoV-2 virus.^30^ Large scale antibody testing should be launched to assess the true scale of this pandemic.^31^ The instrument of isolating nearly everyone with a near universal lockdown not only leads to livelihood losses for millions of families but also starvation for others. As we reopen the country, testing high contact, high density areas and setting up a clever surveillance system is critical.
b. Immediately prepare to protect the health care workers and first responders who are at the front line of this pandemic. This involves ensuring a steady supply chain of medical resources (masks, gloves, gowns, ventilators), and protecting our healthcare workers physically and psychologically. Full gears (protective suit, medical goggle, cap, face shield, mask and gloves) are absolutely essential when seeing suspected cases. These protection strategies worked in China.^32^
c. Reduce all non-essential medical care and expand number of hospital beds, ICU beds and ventilators.
d. Continue to set up COVID-19 testing mobile labs, hospitals and mobile cabins (e.g. by converting stadiums and trains into quarantine and treatment facilities).^33^
e. Ensure the healthcare facilities have adequate supply of medications that are currently being recommended. For instance, antiviral drugs “remdesivir and chloroquine are highly effective in the control of 2019-nCoV infection in vitro” indicating promise for treating COVID-19 patients.^34^ Recently though, the study^35^ finding hydroxycholoroquine as an effective treatment for COVID-19 has been retracted for bad study design and not meeting expected scientific standards.^36^
f. Use pragmatic real-time data for optimally deploying surveillance, community inspection and health care resources. This is key with limited resources.

### Social recommendations

(g) Continue traffic ban, social distancing, quarantine and similar interventions to slow down the spread of the coronavirus even after the lockdown is lifted.
(h) Provide free coverage for testing and treatment for COVID-19 for everyone in India.
(i) From a collective social conscience, COVID-19 patients and caregivers should not be stigmatized or sacrificed.

### Economic recommendations

(j) Provide livelihood assistance over the quarantine period to those who test positive. This will incentivize people to get tested and comply with social isolation protocols. For many people in India, loss of several weeks of earnings can be economically devastating and since symptoms are mild for most infected people, it is unreasonable to expect that all people will tightly follow restrictions unless economically protected. To get a ballpark idea of the fiscal burden involved, assume 1 million detected cases, quarantine of 6 weeks per patient, and INR 10,000 monthly compensation. This adds up to a bill of INR 15 billion, which is roughly 2.5% of the annual healthcare budget of the central government.
(k) During periods of social distancing and lockdowns, there is grave livelihood threat to a lot of poor people even if they are uninfected – street hawkers, auto drivers, barbers and shopkeepers, etc. Providing a universal basic income (UBI), or some mildly means tested version of it, over the period of disruption is needed for a successful lockdown.
(l) To prevent shutdowns in badly affected sectors, the government may provide goods and services tax (GST) credit to firms based on the difference between past and current sales. Once the pandemic is over and normal business resumes, expansionary monetary and fiscal policy will be needed to revive macroeconomic health.

## Role of data science and the troubling existence of many models

There are many epidemiological models to predict the course of an infectious disease and even many that are India-specific.^37–39^ Some use age-structure, contact patterns, spatial information to finesse their prediction. Some consider the possibility of a latent number of true cases, only a fraction of which are ascertained and observed.^16^ The model we used here is an extension of a standard SIR model, called eSIR model,^11^ where we can create hypothetical intervention scenarios in a time dependent manner. The goal of any intervention is to reduce the chance that an infected person meets a susceptible person. We create models for declines/drops in contact probabilities when an intervention is rolled out. Thus, there is some intrinsic ad-hocery to our assumptions. Any statistical model is wrinkled with such assumptions. Similarly, the predictions themselves have large uncertainty (as reflected by the upper credible limits). As we interpret the numbers from any model, let us use caution in not over-interpreting them. A rigorous quantitative treatment often allows us to analyze a problem with clarity and objectivity, but we recommend focusing more on the qualitative takeaway messages from this exercise rather than concentrating on the exact numerical projections or quoting them with certainty.

We see tremendous role of data and data science in governing policy as India reopens post lockdown. The release from lockdown will not be in a binary switch on/switch off process but a modulated slow-varying process. We see the following roles and opportunities for pragmatic use of data science in the post-lockdown phase.

a. Flexible, athletic, data driven policymaking will need up-to-date numbers and projections at hand, which require granular data, automation and data transparency
b. Understand uncertainty in numbers: all models are wrong, some are useful, but note that takeaway messages for intervention forecasting are often the same^40^
c. Using technology to create body temperature/expected health status map (e.g., <u>healthweather.us</u>)^41^
d. Assess adherence to social distancing using mobile networks, Google (e.g., <u>Google Mobility Reports</u>)^42^
e. Use survey to identify potential super-spreaders, manage contact network, oversample high risk areas for testing
f. Install syndromic surveillance in hospitals, medical claims systems to set up alert; establish expected number of respiratory and flu-like illnesses so departures can set off an alarm.
g. Use community health workers in rural areas, community dwellings to identify and isolate cases and conduct cluster testing
h. Targeted communication strategies: regarding tests, treatments, contagion level. Misinformation and incorrectly analyzed data lead to panic
i. Accurate and consistent reporting of case counts and deaths due to COVID-19 are extremely critical

## Limitations

Our statistical modeling and forecasts are not without limitations. We have very few data points and a long time-window to extrapolate for the long-term forecasts. The uncertainty in our predictions is large due to many unknowns arising from model assumptions, population demographics, the number of COVID-19 diagnostic tests administered per day, testing criteria, accuracy of the test results, and heterogeneity in implementation of different government-initiated interventions and community-level protective measures across the country. We have neither accounted for age-structure, contact patterns or spatial information to finesse our predictions^37–39^ nor considered the possibility of a latent number of true cases, only a fraction of which are ascertained and observed.^16^ Increase in frequency and scale of testing, and community transmission of the SARS-CoV-2 virus may lead to a spike in a single day and that can shift the projection curve significantly upwards. COVID-19 hotspots in India are not uniformly spread across the country, and state-level forecasts^43^ may be more meaningful for state-level policy-making. We are assuming that the implementation and effects of public health interventions and policies are the same everywhere in India by treating India as a homogeneous unit. Future opportunities for improving our model include incorporating contagion network, age-structure, estimating SEIR model, incorporating test imperfection, and estimating true fatality/death rates. Regardless of the caveats in our study, our analyses show the impact and necessity of lockdown and of suppressed activity post-lockdown in India. Rather than over-interpreting exact numerical projections, we recommend focusing more on the qualitative takeaway messages.

One ideological limitation of considering only the epidemiological perspective of controlling COVID-19 transmission in our model is the inability to count excess deaths due to other causes during this period, or the flexibility to factor in reduction in mortality/morbidity due to some other infectious or flu-like illnesses, traffic accidents or health benefits of reduced air pollution levels. A more expansive framework of a cost-benefit analysis is needed as we gather more data and build an integrated landscape of population attributable risks.

### Our data science product

Finally, in our strong commitment to reproducibility and dissemination of our research, we have made the code for our predictions available at GitHub (https://github.com/umich-cphds/cov-ind-19) and created an interactive and dynamic R Shiny app (covind19.org) to visualize the observed data and create predictions under hypothetical scenarios with quantification of uncertainties. These forecasts will get updated daily as new data come in. We hope these products will remain our contribution and service as data scientists during this tragic global catastrophe, and the model and methods will be used to analyze data from other countries.

## Conclusion

Our epidemiologic and mathematical calculations make a convincing case for enforcing the 21-day national lockdown of the largest democracy in the world, acting early, before the growth of COVID-19 infections in India starts to accelerate. We also notice the public health benefit of extending the lockdown by 3-5 weeks in our projections. Measures of suppression are needed post-lockdown to get long-term benefits from the lockdown. However, these draconian public health measures come at a tremendous price to social and economic health that can last months or even years after the restrictions on social mobility are lifted. Thus, there is an urgent need for social and economic immunity: not just coverage for testing and treatment for COVID-19 for everyone in India, but subsidies and incentives for the common man to survive the consequences of the severe interventions that are needed to stop the coronavirus from creating a massive catastrophe in India. We also illustrate the critical role of data in aiding policy decisions.

Finally, our message to the public is to proceed with prudence and caution, and not panic or drown in despair. We should draw hope from the success of South Korea and China and the initial promising containment in India. We need to support the community around us and help the government of India to manage the crisis with the best strategies, resources and science. The lockdown has given us time to prepare and act, let us make the best use of it. We are still in a state of national and global emergency and it will take a considerable time for humanity to recover from this global pandemic and return to normalcy. In the meantime, we root for **public health**, for **innovation and science**, for home testing kits [<u>there is none yet</u>],^44^ for FDA approved drugs [<u>Solidarity Trial</u>],^45^ and for a vaccine [<u>5 clinical trials ongoing</u>].^46^ In these frightening times, we find inspiration in the power of the common people and the magic of human kindness.

## Data Availability

All data and prediction codes used for analysis in this article are publicly available and are detailed in the article.

http://covind19.org/

## Acknowledgement

The authors will like to thank the University of Michigan Advanced Research Computing Services for enabling daily updates to our models and allocating us abundant computational resources. We will also like to thank Professor Matthew Fox from the Boston University School of Public Health for his valuable comments on our RShiny App,

---

**Supplementary Table 1.** Proportion of population in specifically vulnerable subgroups at potentially high risk of COVID-19 severity risk in India.

| **Metric** | **Number<sup>†</sup>**<br>**(in millions)** | **Year** | **Source** |
| --- | --- | --- | --- |
| Uninsured | 1,104 | 2014 | Prinja et al. 2019 |
| Population over 65 | 92.5 | 2020 (est.) | CIA World Factbook |
| Hypertension (men)* | 175.7 | 2015/16 | Gupta & Ram 2019 |
| Hypertension (women)* | 132.6 | 2015/16 | Gupta & Ram 2019 |
| People with cardiovascular disease* | 78.7 | 2016 | Prabhakaran et al. 2018 |
| Population with COPD* | 75.9 | 2016 | Lancet 2018 |
| Population with asthma* | 45.5 | 2016 | Lancet 2018 |
| Develop cancer by age 75 (men)** | 70.3 | 2018 | NICPR |
| Develop cancer by age 75 (men)** | 62.3 | 2018 | NICPR |
| Population with diabetes (adult) | 122.8 | - | IDF |
| Access to inpatient department facilities*** | 731.4 | 2012 | IMS Institute 2013 |
| Access to outpatient department*** | 1,104 | 2012 | IMS Institute 2013 |

<sup>†</sup> based on 2020 est. of 1.38 billion from UN Department of Economic and Social Affairs
\* age-standardized; \*\* risk; \*\*\* defined as within 5-kilometer distance of home or work
Abbrev.: COPD, chronic obstructive pulmonary disease; IDF, International Diabetes Federation; NICPR, National Institute of Cancer Prevention and Research

**Supplementary Table 2.** Intervention landscape of countries severely affected by COVID-19

| Country | Date of 1st case<sup>†</sup> | Interventions/Lockdown | Crude fatality rate<sup>†</sup> | Active cases<sup>†</sup> |
| --- | --- | --- | --- | --- |
| China | Nov 17, 2019* | Lockdown in Wuhan on 01/22, extended to neighboring cities in Hubei province on 01/23. Wuhan lockdown to be lifted on 04/08.<sup>1</sup> | 4.1% | 1,116 |
| South Korea | Jan 19, 2020** | Tested widely for the virus, isolated cases and quarantined suspected cases. Figures indicate that this has helped suppress transmission of the virus. The country appears to have reined in the outbreak without some of the strict lockdown strategies deployed elsewhere in the world.<sup>2</sup> | 2.0% | 3,125 |
| United States | Jan 20, 2020 | Onn 01/31, restricted travel from China, expanded restrictions to other countries on 02/29. On 03/03 CDC lifted all restrictions on testing. On 03/15, CDC recommends no gatherings of 50 people or more. Stay-at-home directives issued at state-level.<sup>3</sup> | 3.7% | 431,557 |
| France | Jan 24, 2020 | On 03/17, France imposed a nationwide lockdown, prohibiting gatherings of any size and postponing the second round its municipal elections. The lockdown was one of Europe's most stringent. While residents were told to stay home, officials allowed people to go out for fresh air but warned that meeting a friend on the street or in a park would be punishable with a fine.<sup>3</sup> | 10.8% | 79,279 |
| Germany | Jan 28, 2020 | Germany has a National Pandemic Plan, with three stages. In the first stage (containment) health authorities are focusing on identifying contact persons, who are put in personal quarantine and are monitored and tested. In the second stage (protection) the strategy will change to using measures to protect vulnerable persons from becoming infected. The final stage (mitigation) will try to avoid spikes of intensive treatment in order to maintain medical services.<sup>4</sup> | 2.2% | 76,420 |
| United Kingdom | Jan 31, 2020 | Citizens advised to stay at home. Violators will be fined with the exception of few special circumstances, in order to contain the spread of the disease. The government is supporting and coordinating with research institutes to explore treatment and curative options.<sup>5</sup> | 12.7% | 61,825 |
| Italy | Jan 31, 2020 | Flights to China suspended and a national emergency declared on 01/31 after two cases are confirmed in Rome. Schools and universities closed on 03/04. By 03/09, the entire nation placed under lockdown, with restaurants, bars closing on 03/11 and factories closing on 03/22. All non-essential production halted.<sup>6</sup> | 12.7% | 96,877 |
| Spain | Feb 1, 2020 | State of emergency declared on 03/14, allowing authorities to authorities to confine infected people and ration goods. Originally planned to last till 03/29, has been extended to 04/12. Schools, bars, restaurants and shops selling non-essential items have been shut since March 14 and most of the population is house bound.<sup>7</sup> | 10.2% | 85,386 |
| Iran | Feb 19, 2020 | On 02/28, the Iranian authorities closed schools, canceled Friday prayers and moved to restrict visitors from China. On 03/15, the official leading Iran's response to the new coronavirus acknowledged Sunday that the pandemic could overwhelm health facilities in his country, which is battling the worst outbreak in the Middle East.<sup>8</sup> | 6.2% | 28,495 |

¶ *Date of 1<sup>st</sup> case data obtained from JHU CSSE time series data on COVID-19 (except for China & South Korea)*
† Microsoft Bing COVID-19 tracker (https://bing.com/covid, as of 1:00 PM EST April 10, 2020)
\* https://www.livescience.com/first-case-coronavirus-found.html
\*\* https://www.worldometers.info/coronavirus/
1 https://www.cnn.com/2020/03/24/asia/coronavirus-wuhan-lockdown-lifted-intl-hnk/index.html
2 https://www.npr.org/sections/goatsandsoda/2020/03/26/821688981/how-south-korea-reigned-in-the-outbreak-without-shutting-everything-down
3 https://www.nytimes.com/article/coronavirus-timeline.html
4 https://en.wikipedia.org/wiki/2020_coronavirus_pandemic_in_Germany
5 https://www.gov.uk/government/publications/coronavirus-action-plan/coronavirus-action-plan-a-guide-to-what-you-can-expect-across-the-uk
6 https://www.axios.com/italy-coronavirus-timeline-lockdown-deaths-cases-2adb0fc7-6ab5-4b7c-9a55-bc6897494dc6.html
7 https://www.reuters.com/article/us-health-coronavirus-spain/shellshocked-spain-reports-record-832-new-coronavirus-deaths-idUSKBN21F0NR
8 https://www.nytimes.com/aponline/2020/03/15/world/middleeast/ap-m1-virus-outbreak-mideast.html?searchResultPosition=10

**Supplementary Figure 1.**
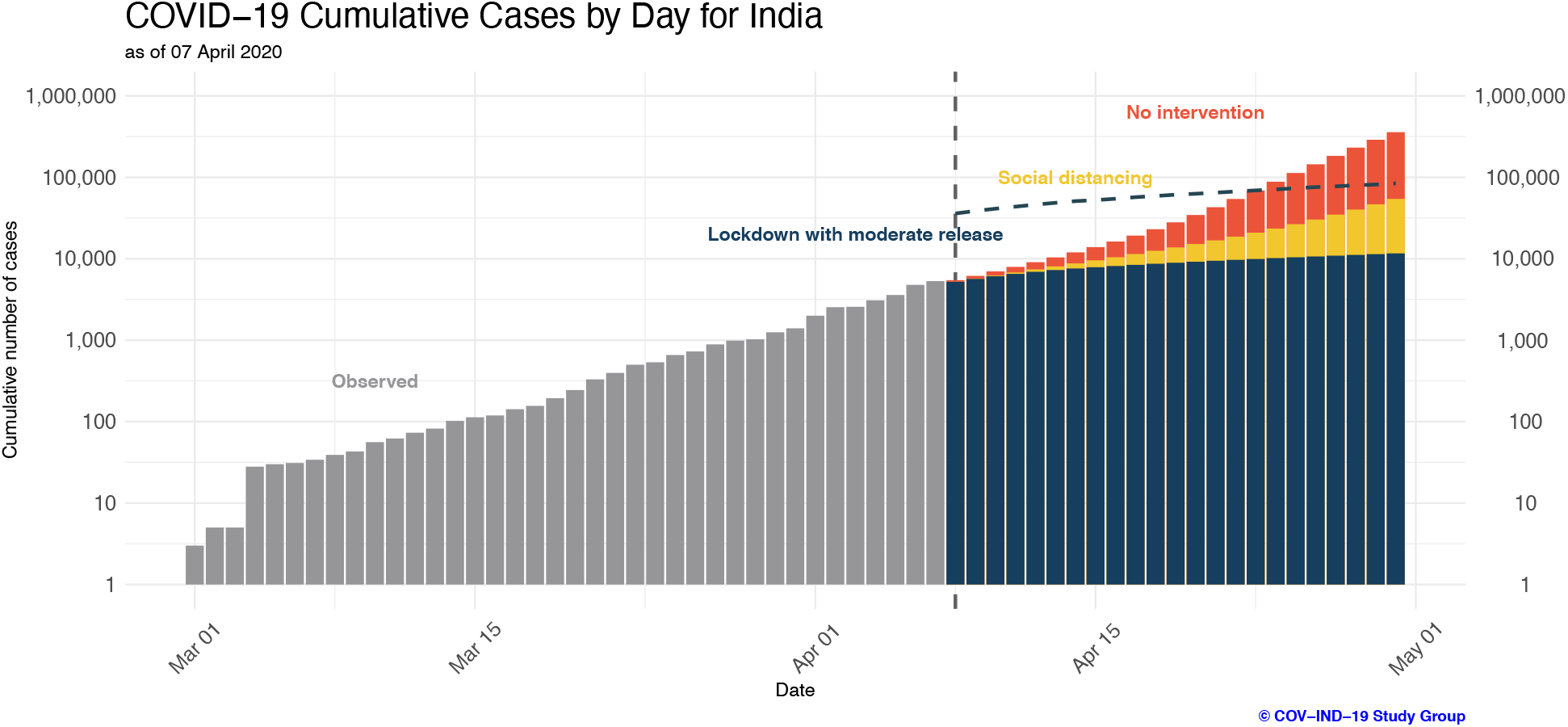
Short-term daily growth in cumulative case counts in India assuming a 2-week delay in people’s adherence to restrictions. Observed data are shown for days up to April 7. Predicted future case counts for April 8 until April 30 are based on observed data until April 7 using the eSIR model.

**Supplementary Figure 2.**
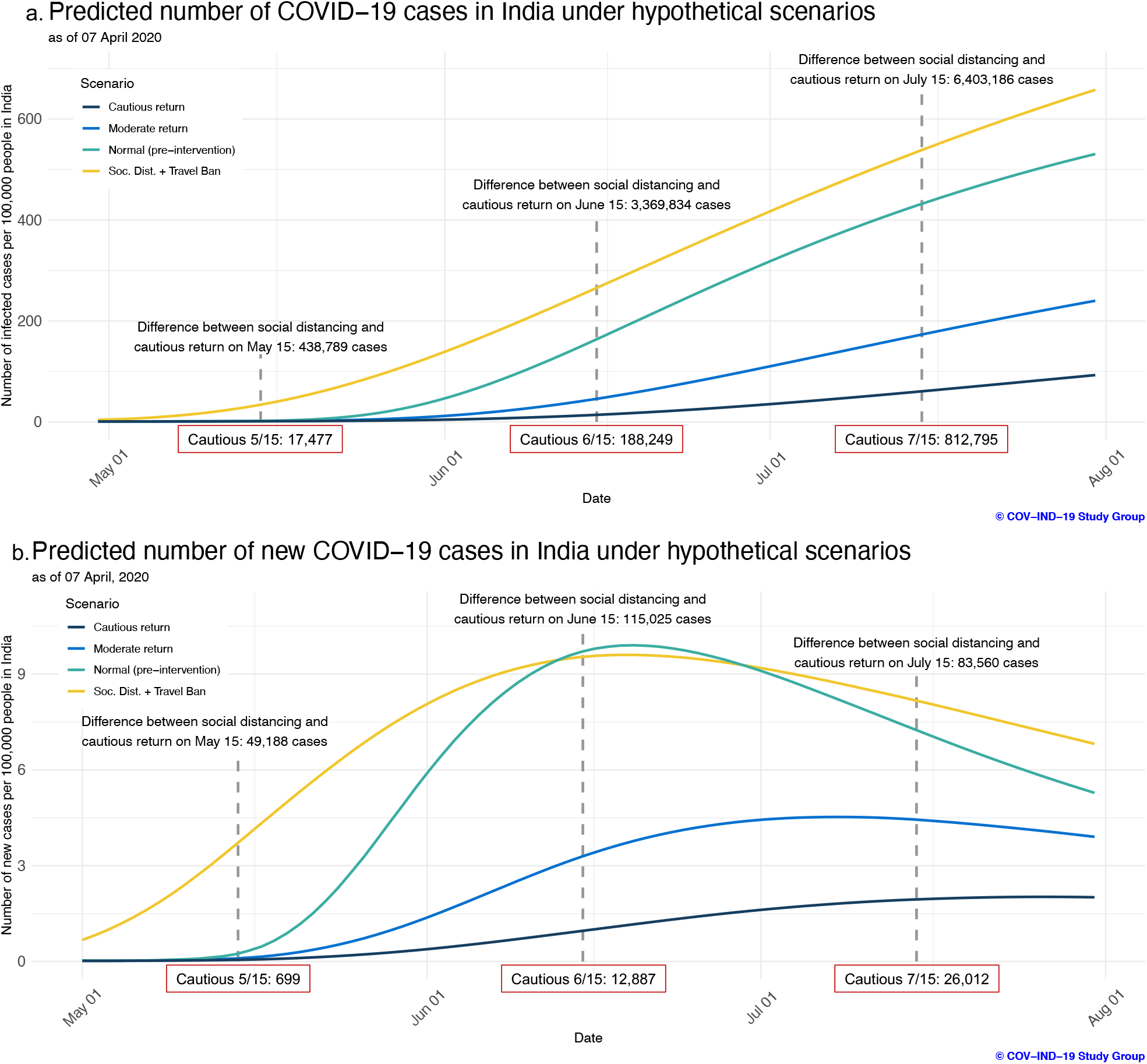
Long-term daily growth in case counts in India per 100,000 people assuming a 2-week delay and how that is affected by different non-pharmaceutical intervention strategies. Predicted cumulative (a) and incident (b) case counts from April 30 to July 31 from the eSIR model are shown, based on observed data until April 7.

**Supplementary Figure 3.**
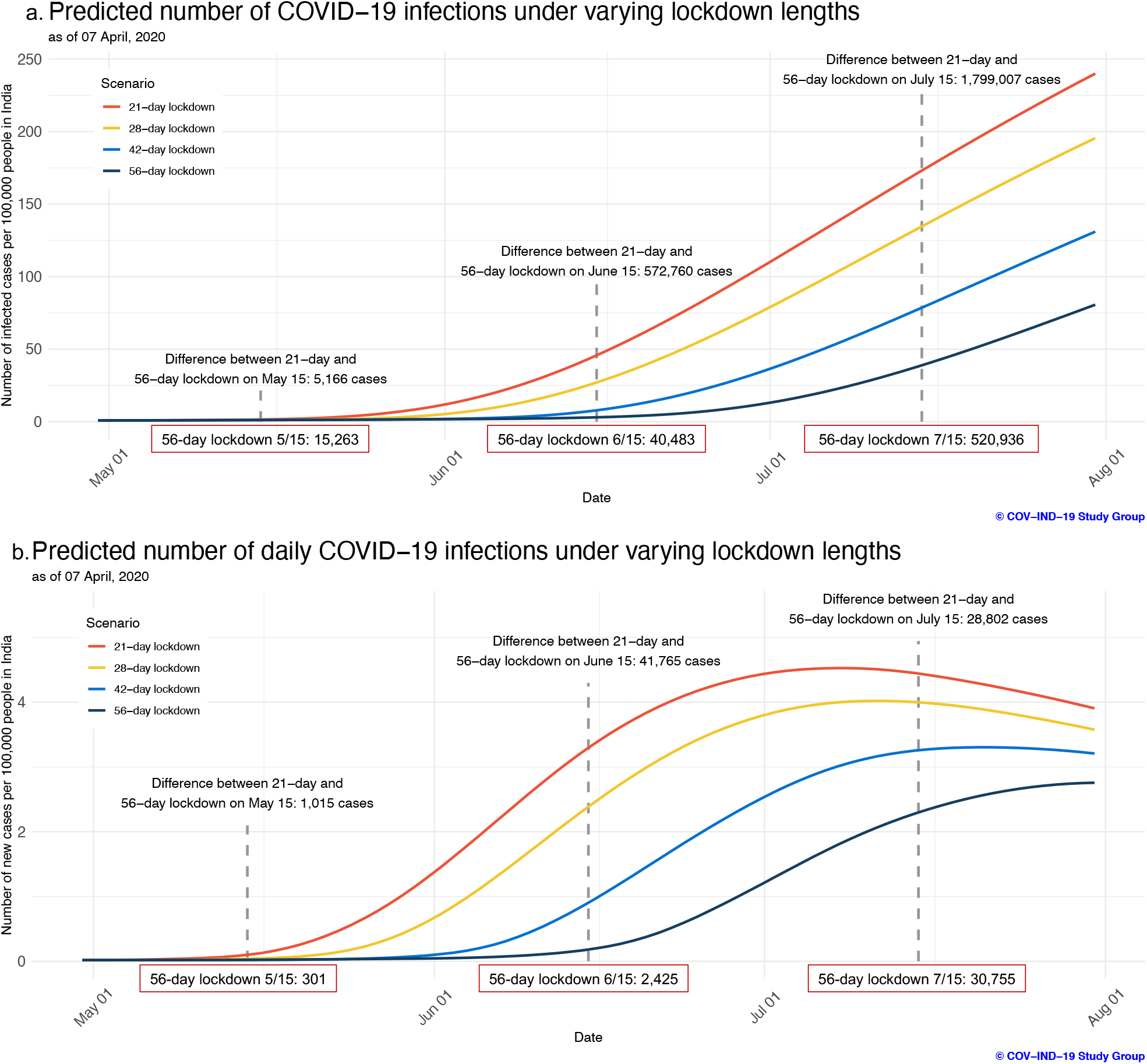
Cumulative (a) and incidence (b) graphs for forecasting models assuming a 2-week delay under 21-, 28-, 42-, and 56-day lockdown scenarios using observed data through April 7.

**Supplementary Figure 4.**
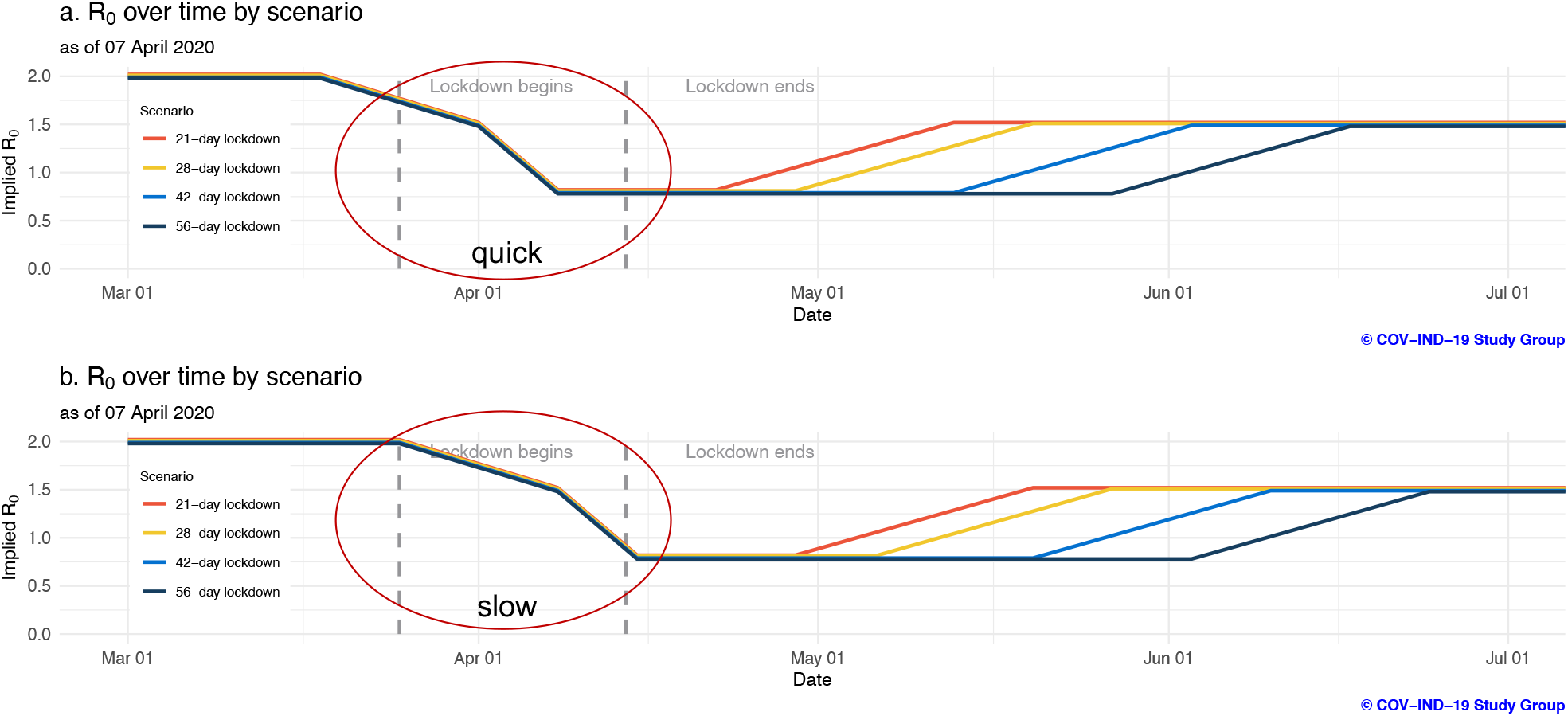
Implied R_0_ schedules corresponding to quick and slow adherence for the hypothetical lockdown duration scenarios.

